# Human liver organoid derived intra-hepatic bile duct cells support SARS-CoV-2 infection and replication and its comparison with SARS-CoV

**DOI:** 10.1101/2021.02.10.21251458

**Authors:** Vincent Chi-Hang Lui, Kenrie Pui-Yan Hui, Rosanna Ottakandathil Babu, Haibing Yue, Patrick Ho-Yu Chung, Paul Kwong-Hang Tam, Michael Chi-Wai Chan, Kenneth Kak-Yuen Wong

## Abstract

**Background:** Although the main route of infection for severe acute respiratory syndrome coronavirus 2 (SARS-CoV-2) is the respiratory tract, liver injury is also commonly seen in many patients, as evidenced by deranged parenchymal liver enzymes. Furthermore, patients with severe liver disease have been shown to have higher mortality. Overall, the mechanism behind the liver injury remains unclear.

**Approach and results:** We showed that intra-hepatic bile duct cells could be grown using a human liver organoid platform. The cholangiocytes were not only susceptible to SARS-CoV-2 infection, they also supported efficient viral replication. We also showed that SARS-CoV-2 replication was much higher than SARS-CoV.

**Conclusion:** Our findings suggested direct cytopathic viral damage being a mechanism for SARS-CoV-2 liver injury.

## Introduction

Coronavirus disease-2019 (COVID-19) is caused by severe acute respiratory syndrome coronavirus 2 (SARS-CoV-2) and has rapidly become a worldwide pandemic. As of January 2021, there have been 83,322,449 confirmed cases of COVID-19 and 1,831,412 deaths worldwide [1].

SARS-CoV-2 is a single-strand positive-sense RNA virus, belongs to the beta coronavirus family, which enters cells through the Angiotensin Converting Enzyme 2 (ACE2) receptor [2]. SARS-CoV-2 interacts with the host cells by first attaching its spike protein to ACE2 on host cells, and then gains entry through hemagglutinin cleavage by host cell protease Transmembrane protease, serine 2 (TMPRSS2) [3, 4]. Human-to-human transmission for SARS-CoV-2 is efficient. While the respiratory tract is a common route of disease transmission [5, 6], the gastrointestinal tract has also been shown as another possible route of viral transmission [7-9]. Understanding the cellular/tissue tropism and the route of infection of SARS-CoV-2 virus is essential in overall patient management and infection control.

Liver damage is often identified as a typical occurrence in COVID-19 patients, and 58%–78% of COVID-19 patients were shown to exhibit various degrees of liver injury [10]. Some COVID-19 patients have elevated levels of liver enzymes such as aspartate amino-transferase (AST), alanine aminotransferase (ALT) levels, and gamma-glutamyl transferase (GGT), while some patients have higher overall bilirubin levels and lower serum albumin [11-13]. Indeed, elevated AST, ALT, and total bilirubin levels but lower serum albumin levels are correlated with higher death rate [6], and have been found in the severe group of COVID-19 patients [14]. Moreover, activation of coagulation and fibrinolysis accompanied by thrombocytopenia was observed in severe COVID-19 cases [15]. Autopsy examinations of a small number of COVID-19 patients have provided conclusive evidence of secondary liver injury [16, 17]. Liver damage can be aggravated by the increase of COVID-19 infection severity, which indicates that the degree of liver damage may serve as an indicator of COVID-19 progression. Nonetheless, the mechanism of liver injury is poorly understood and may be due to direct viral hepatitis, bystander systemic inflammatory response or complications of drug treatment.

Angiotensin converting enzyme 2 (ACE2), the protein through which SARS-CoV-2 gains entry, is abundantly expressed on many cells, including liver cells, bile duct cells and liver endothelial cells. ACE2 expression levels in bile duct cells is slightly higher than those in hepatocytes and is comparable with alveolar epithelial type II cells [18]. Given that bile duct cells play an important role in immune defense and liver regeneration, their impairment may serve as a major cause of virus-induced liver injury in COVID-19 patients [19]. Although typical coronavirus particles characterized by spiked structures in the cytoplasm of hepatocytes have been identified [20], the susceptibility of bile duct cells to SARS-CoV-2 infection is yet to be confirmed.

An organoid is a miniaturized and simplified version of an organ produced *in vitro* in 3-dimension. It shows realistic micro-anatomy and retain the biology of individual tissues. Lung and gut organoids have been successfully used to demonstrate SARS-CoV-2 infection [21, 22]. Recently, human liver ductal organoids were shown to express ACE2 and TMPRSS2, and were permissive to SARS-CoV-2 infection [23]. The extra-and intra-hepatic bile duct cells are derivatives of different progenitors, in that extra-hepatic ducts arise from a common SOX17+/PDX1+ pancreatobiliary progenitor, while the intra-hepatic ducts arise from CK19+/AFP+ hepatoblasts [24]. Furthermore, the cellular properties and functions of extra- and intra-hepatic cholangiocytes (bile duct cells) are very different. It remains to be investigated if intra-hepatic bile ducts are susceptible to SARS-CoV-2 infection and support viral replication. Recently, we have shown that the human liver tissue organoids derived from EPCAM+ve cells of liver biopsies are from the hepatoblast progenitor (CK19+) lineage rather than from the pancreatobiliary (extrahepatic ducts) progenitor (SOX17+/PDX1+) lineage, and thus recapitulates the intrahepatic cholangiocyte development [25].

In this study, we utilized human liver tissue derived organoids from hepatoblast progenitor as an ex-vivo tool to investigate the infection, tropism and replication competence of SARS-CoV-2 on intra-hepatic bile ducts and compared to SARS-CoV. We showed that liver tissue derived organoids consisted of intra-hepatic cholangiocytes and hepatoblasts as major cell types, which expressed ACE2 and TMPRSS2 and could be rapidly infected by both SARS-CoV-2 and SARS-CoV. SARS-CoV-2 was shown to have a much higher replication rate than SARS-CoV.

## Materials & Methods

### Liver tissues

Pediatric liver tissues were obtained from non-tumour margin of hepatoblastoma (HB). Liver biopsies were obtained during operations with full informed consent from parents or patients, and the study was approved by Hong Kong West Cluster-Hong Kong University Cluster Research Ethics Committee/Institutional Review Board (UW 16-052).

### Human pediatric liver organoid culture

Wedge liver biopsies (1-2 cm^3^) from the non-tumor margin of pediatric patients (n=4) with hepatoblastoma (HB)). Liver tissues were minced in cold wash medium (Advanced DMEM/F12; 1% GlutaMAX; 1% FBS; 1% Penicillin/Streptomycin (P/S)) and digested in digestion medium (5 ml, Multi Tissue Dissociation Kit 1; Miltenyi Biotec Inc. CA, USA) in a gentleMACS-C Tube (Miltenyi Biotec Inc. CA, USA) on gentleMACS™ Octo Dissociator (Miltenyi Biotec Inc. CA, USA) using program 37°C-Multi-A-01. After digestion and washing with cold wash medium (5 ml), the digested sample was passed through a 70 µm strainer and then through a 30 µm strainer before centrifugation (300 g; 10 min) to pellet the cells. Cell pellet was resuspended cells in 200 µl of MACS column buffer (PBS pH 7.2 + 0.5 % BSA), and incubated with CD326 (EPCAM) microbeads (1 µl microbeads/2×10^6^ cells; 130-090-500; Miltenyi Biotec Inc. CA, USA) in the dark at 4°C for 30 minutes. After incubation, column buffer (0.5 ml) was added and EpCAM positive cells were sorted on MS Column following the manufacturer’s protocol (Miltenyi Biotec Inc. CA, USA). The cells were pelleted (300 g; 10 min.) and resuspended in 200 µl of column buffer for cell counting. The cell pellet (approximately 1×10^5^ cells) was mixed with 50-60 µl of Matrigel (356231; Corning Biocoat) and seeded per well of a prewarmed (37°C) 4-well plate (176740; Nunc; Thermo Scientific). After Matrigel had solidified, organoid medium (500 µl) was added to each well. Organoid medium was based on Advanced DMEM/F12 (Invitrogen) supplemented with Penicillin/Streptomycin (Invitrogen), GlutaMax (Invitrogen), 25 mM HEPES (Invitrogen), 1% N2 (GIBCO), 1% B27 (GIBCO), 1.25 mM Acetylcysteine (Sigma), 10 nM gastrin (G9145; Sigma), 50 ng/ml EGF (PMG8043; PeproTech), 100 ng/ml FGF10 (100-26-25UG; PeproTech), 25 ng/ml HGF (100-39-10UG; PeproTech), 10 mM Nicotinamide (Sigma), 5 µM A83.01 (Tocris), 10 µM Forskolin (Tocris), 500 ng/ml R-Spondin 1 (7150-RS-025; R&D), 100 ng/ml Noggin (250-38-20UG; Peprotech), 100 ng/ml Wnt3a (1324-WN-010; R&D) and 250 ng/ml Amphotericin B (#15290018; GIBCO). For the first six days of culture, 10 µM ROCK inhibitor Y-27632 (Tocris) was added. Medium was changed once every two days.

### Single cell RNA Sequencing and data analysis

Single cells from organoids were prepared. The organoid medium along with matrix gel containing organoids was transferred to a 15 ml tube, 1-2 ml cold Advanced DMEM/F12 was added and the mixture was incubated on ice for 10 min to dissolve matrix gel before centrifugation (300g; 5 min). The supernatant was aspirated until the organoid pellet and a layer of matrix gel remained. 1 ml 5X TrypLE Express (12604013; GIBCO) was added, mixed well and incubated at 37°C for 5 min. 1 ml FBS was added to the mixture, which was pipetted up and down for 40-50 times with a small circumference opening glass pipette (diameter 0.3-0.5 mm) to efficiently dissociate organoids into single cells. 5-10 ml cold Advanced DMEM/F12 medium was added, and the suspension was passed through a 30 µm strainer before centrifugation (300 g; 5 min) at 4°C. The supernatant was aspirated until only the pellet remained and the cells were counted following addition of 200 µl 10% FBS. The cells were then counted using Invitrogen Countess II FL Automated Cell Counter. Samples were prepared as outlined by the 10x Genomics Single Cell 3’ v2 Reagent Kit user guide. The samples with more than 1 x 10^6^ cells/ml along with >80% viability is then resuspended in 50-100ul, 10% FBS and are submitted for 10X Genomics single-cell sequencing at Centre for PanorOmic Sciences (CPOS)-HKU. Sample viability was double confirmed by Trypan Blue (Thermo Fisher) and using a haemocytometer (Thermo Fisher). Following counting, the appropriate volume, ∼33.8ul of each sample were used for cell encapsulation for a target capture of 4000 cells. Sequencing run were carried out using Illumina NovaSeq 6000 for those sequencing libraries passing the QC with good cDNA yield. Single cell transcriptomic data have been uploaded to NCBI Sequence Read Archive (SRA) and can be found with accession number: PRJNA609259.

The CellRanger (10X Genomics) analysis pipeline was used to generate digital gene expression matrix (UMI counts per gene per cell) from sequencing data by aligning to the human genome. The raw gene expression matrix was filtered, normalized and clustered using standard Seurat package procedures 16. The low quality cells were removed from the analysis using the following thresholds: cells with a very small library size or UMI counts per cell (nUMI >1500), genes detected per cell (nGene >1000), UMIs vs. genes detected (log10GenesPerUMI >0.8), mitochondrial counts ratio (mitoRatio <0.1). Cell-cycle phases were predicted using a function included in Seurat that scores each cell based on expression of canonical marker genes for S and G2/M phases, this was used to regress out the cell-cycle effects from the downstream analysis. Clusters were visualized using uniform manifold approximation and projection (UMAP) as implemented in Seurat. First 13 PCs were selected for UMAP based on the points where the principal components cumulatively contribute 90% of variation associated with entire data. The cell-type identities for each cluster in UMAP were determined using presence of known marker genes in each cluster.

### Virus stock preparation

A SARS-CoV-2 (BetaCoV/Hong Kong/VM20001061/2020, SCoV2) was isolated from a COVID-19 patient in Hong Kong in 2020. For comparison, we used a SARS-CoV (strain HK39849, SCoV) isolated from a hospitalized SARS patient in Hong Kong in 2003. SARS-CoV-2 and SARS-CoV viruses were propagated in Vero-E6 cells and virus stock was titrated to determine tissue culture infection dose 50% (TCID_50_) in Vero-E6 cells. The experiments were carried out in a Bio-safety level 3 (BSL-3) facility.

### Coronavirus infection

The 3D liver organoids were sheared mechanically using syringe to expose the apical surface to the virus inoculum. Around 100-200 organoids were infected with each coronavirus at 5×10^5^ TCID_50_/ml for 1 h at 37°C. The organoids were washed three times with culture medium, re-embedded in Matrigel at the same conditions with the same growth medium and incubated at 37°C with 5% CO_2_. The viral titers in the culture supernatants were measured at 1, 24, 48, and 72 h after infection using the TCID_50_ assay in Vero-E6 cells. Cell lysates were collected 72h after infection to assess mRNA expression of cytokines. Organoids were fixed 72 h after infection in paraformaldehyde for immunofluorescent staining.

### Immunofluorescence

Organoids in matrigel (from 2 HB livers) were fixed in 4% paraformaldehyde (w/v) in PBS (phosphate-buffered saline, pH 7.2) for 48 h at 4°C, dehydrated in graded series of alcohol, and cleared in xylene before being embedded in paraffin. Sections (6 µm in thickness) were prepared and mounted onto TESPA-coated microscope glass. Sections were dewaxed in xylene, hydrated in a graded series of alcohol and finally in distilled water. Antigen retrieval was performed by incubation of slides in Citrate buffer (pH 6.0) at 95°C for 10 minutes. After blocking in PBS-T (PBS with 0.1% Triton) supplemented with 1% Bovine Serum Albumin for 1 h at room temperature, sections were incubated with antibody diluted in PBS-T/BSA for overnight at 4°C. After washing in PBS-T, sections were incubated with fluorescent tagged secondary antibodies in PBS-T/BSA at 37°C for 1 h. Details of the primary and secondary antibodies and their dilutions are shown in Table 1 (supplementary data). After PBS-T washings, sections were mounted in DAPI-containing anti-fade mounting fluid. Images were taken with Nikon Eclipse 80i microscope mounted with a SPOT RT3 microscope digital camera under fluorescence illumination. Photos were compiled using Adobe Photoshop CS6.

### Quantitative RT-PCR

The RNA of infected cells was extracted at 72 h post infection using a MiniBEST universal RNA extraction kit (Takara Biotechnology). RNA was reverse-transcribed by using oligo-dT primers with RT-PCR kit (Takara). mRNA expression of target genes was performed using an ABI ViiA™7 real-time PCR system (Applied Biosystems). The gene expression profiles of cytokines and chemokines were quantified and normalized with β-actin as previously described [26].

### Statistical analysis

Statistical analysis was done using GraphPad Prism software version 9. Experiments with the human organoids were performed independently in two different donors each with triplicate wells. Viral titers and area under the curve (AUC) derived from viral titers and mRNA expression were compared using one- or two-way ANOVA with Tukey’s multiple comparisons test. Mock infected organoids served as negative controls. Results shown in figures are the calculated mean and standard deviation of mean. Differences were considered significant at *p* < 0.05.

## Results

### Expression of ACE2 and TMPRSS2 in human liver organoids

To investigate if human liver derived organoids can be used to establish an *ex vivo* SARS-CoV-2 infection model for intra-hepatic bile ducts, we first determined whether the organoid culture could preserve the cholangiocytes expressing ACE2 and TMPRSS2 *ex vivo*. We performed single-cell RNA sequencing (scRNA-seq) analysis of human liver organoids from 2 patients to interrogate the transcriptomic signatures of cells in human liver organoids. The organoids used in the study are derived from the hepatoblast (intrahepatic ducts) progenitor (CK19+) lineage, and thus recapitulate the intrahepatic cholangiocyte development. A total number of 5885 cells were analyzed and cell populations were visualized by uniform manifold approximation and projection (UMAP), partitioning the cells into three clusters (Figure 1). The common cholangiocyte markers epithelial cell adhesion molecule (EPCAM) and keratin 19 (CK19) were uniformly highly expressed in all the clusters, indicating the heterogeneity of cholangiocytes in these organoids was relatively low (Figure 1). Notably, we identified the SARS-CoV-2 receptor gene ACE2 expressed sparsely among all the cluster in unbiased preferences and was detectable in 7.32% cells of developing cholangiocytes, 4.78% cells in cholangiocytes and 17.48% cells in hepatoblasts (Figure 1B and 1C). Besides, TMPRSS2 expressed uniformly across all the clusters and accounted for 30.18% of developing cholangiocytes, 20.52% in cholangiocytes and 17.89% in hepatoblasts, it is worth mentioning that the ACE2+ cells were co-expressing TMPRSS2 (159 out of 233) (Figure 1B and 1C), making this cell population potentially highly vulnerable to SARS-CoV-2 infection. Interestingly, these same cells were also found to express another serine protease, TMPRSS4. Immunostaining further verified the presence of ACE2+, TMPRSS2+ cholangiocytes (CK19+) in human liver ductal organoids (Figure 1D). Taken together, our data demonstrate that human liver tissue derived organoid preserves the human-specific ACE2+/TMPRSS2+ population of intra-hepatic cholangiocytes.

**Figure 1.**
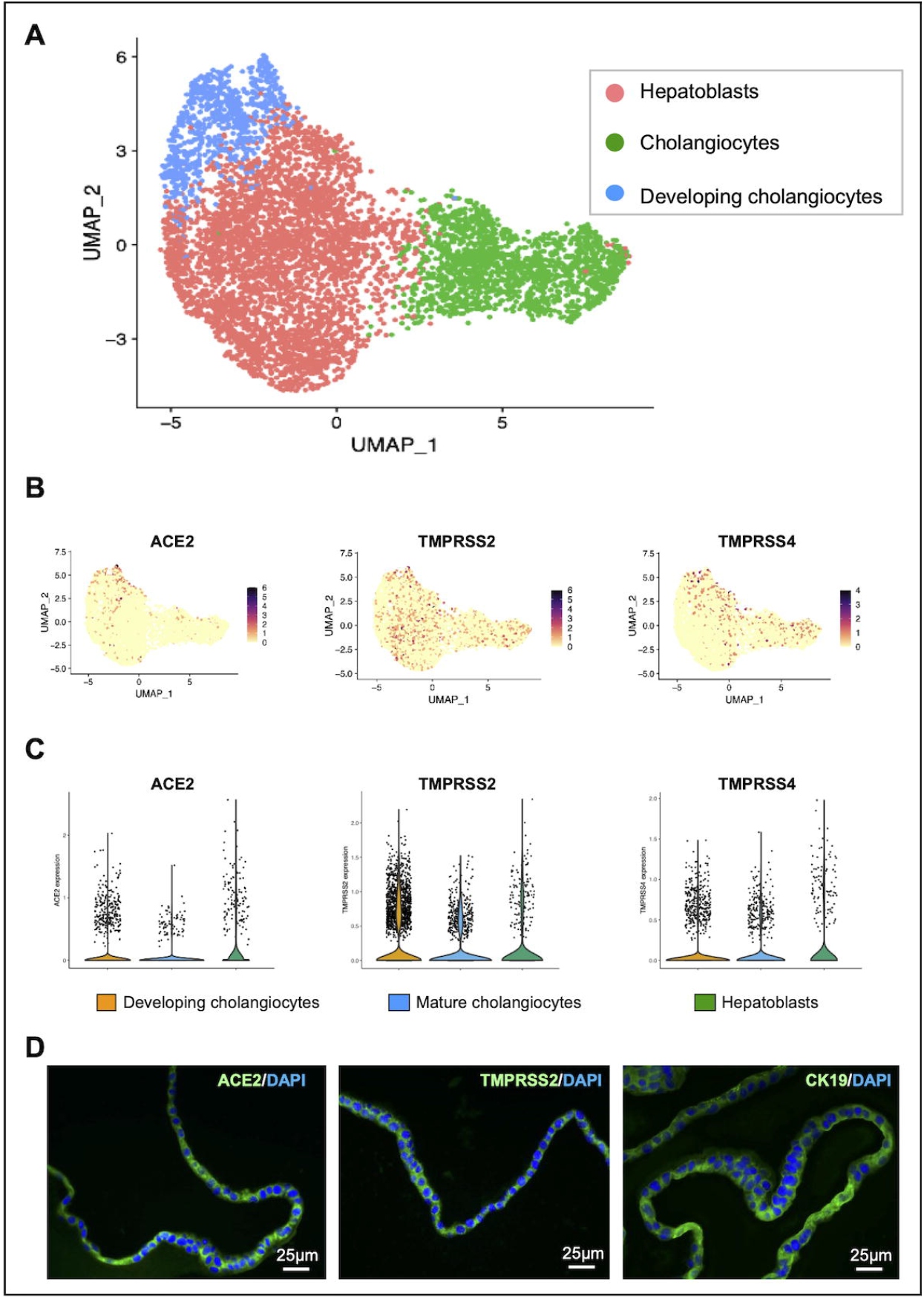
(A) UMAP showing the assigned identity for each cluster (3 clusters identified) in 10X Genomics single-cell RNA sequencing analysis of human control liver organoids. (B & C) Feature plot and violin plot displaying single cell expression distributions for features (ACE2, TMPRSS2 and TMPRSS4) in each cluster. (E) Immunofluorescence staining of human liver organoids for ACE2, TMPRSS2 and TMPRSS4.

### Intra-hepatic cholangiocytes are susceptible to SARS-CoV-2 infection

Next, we examined the susceptibility of human liver ductal organoids to SARS-CoV-2. We infected liver organoids with the SARS-CoV-2, and the SARS-CoV. The liver organoids from two individuals were inoculated with viruses for 1 h, re-embedded in Matrigel and cultured for 48 and 72 hrs. We performed immunostaining to identify the virus-positive cholangiocytes 48 h and 72 h post-infection (Figure 2 and data not shown). The expression of SARS-CoV nucleocapsid protein (Sco-Np) was detected in patchy areas of human liver organoids infected with SARS-CoV-2 and SARS-CoV, whereas no signal was found in uninfected control (Mock, Figure 2A).

**Figure 2.**
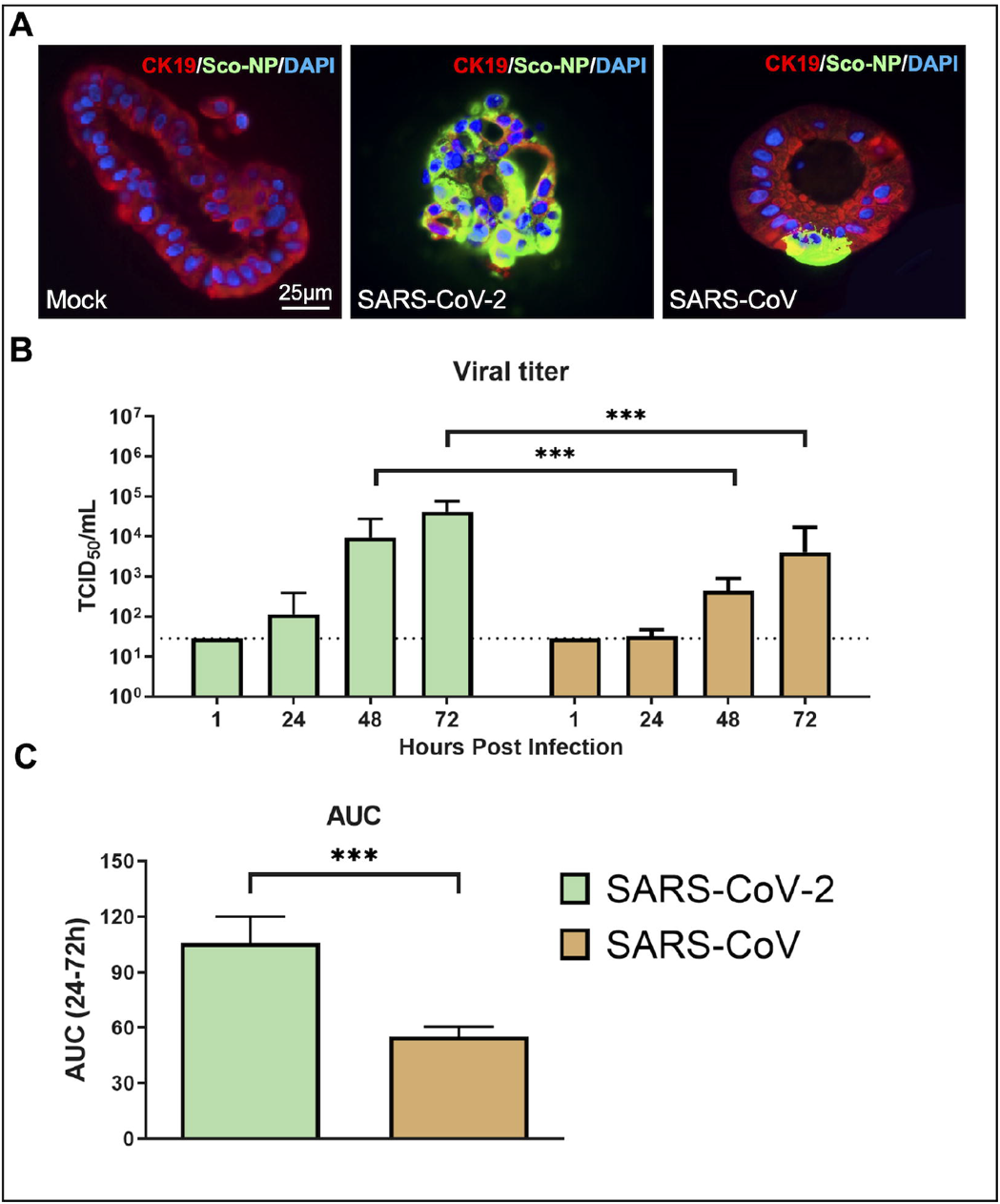
Viral infection and replication kinetics of SARS-CoV-2 and SARS-CoV viruses in human liver organoids. (A) Immunofluorescence staining for SARS-CoV nucleocapsid protein (Sco-Np) of mock and viral infected organoids at 48 h post-infection. (B) Bar chart showing viral titres in culture supernatants of human liver organoids infected with SARS-CoV-2 and SARS-CoV viruses. The organoids were infected with 5×10^5^ TCID_50_/mL at 37°C. Culture supernatants were harvested at the indicted times and virus titres were measured by TCID_50_ assay. Bar-charts show the mean virus titre ± standard deviation (SD). The horizontal dotted line denotes the limit of detection in the TCID_50_ assay. Statistical significances compared among viruses were calculated using two-way ANOVA with Tukey’s multiple comparisons test. \*\*\**p*<0·001. (C) Bar-charts showing the area under the curve (AUC) derived from the viral titres from 24 h to 72 h. Data are presented as mean ± standard deviation (SD). Statistical significances compared among viruses were calculated using one-way ANOVA with Tukey’s multiple comparisons test. \*\*\**p*<0·001.

### Intra-hepatic cholangiocytes supports robust replication of SARS-CoV-2

Virus tropism and replication competence of SARS-CoV-2 with SARS-CoV were compared. Here, we showed that the viral titres of SARS-CoV-2 increased starting at 24h and with more than 2 log increase from 24h to 72h in bile duct organoids. SARS-CoV-2 replicated significantly higher than that of the SARS-CoV, with about 1 log increase in viral titer along at all-time points (Figure 2B). When comparing the area under the curve (AUC) derived from the viral titers from 24h to 72h, similar observations were found, in that SARS-CoV-2 had a significantly higher viral replication competence than SARS-CoV (Figure 2C). These data demonstrate that human bile duct organoids are susceptible to SARS-CoV-2 infection and support robust viral replication.

## Discussion

COVID-19, which is caused by SARS-CoV-2, has become a significant global pandemic since January 2020. Now one year on, there have been over 70,000,000 confirmed cases and 2,000,000 deaths worldwide [1]. Although the main route of SARS-CoV□2 infection remains the respiratory tract, with infected patients displaying symptoms of severe respiratory compromise, the digestive system has been reported to be another potential portal of viral infection [7-9, 21]. Indeed, ACE2 has now been shown to be abundantly expressed in intestinal cells and SARS-CoV-2 can invade and enter via the gastrointestinal tract [27,28].

In many COVID-19 patients, liver damage, as typified by deranged liver function tests, is seen during the SARS-CoV-2 infection [10,11]. Despite this, the mechanisms of liver injury remain largely undetermined. It has been postulated that direct viral infection, drug cytotoxicity, and a bystander inflammatory immune response may play a role. Given the fact that ACE2 has also been confirmed to be present on liver cells [29] and that the liver is connected to the gastrointestinal system via the biliary tract, it is indeed a possibility that the liver may also be a potential target for SARS-CoV-2 infection. Previous report on the detection of SARS□CoV in liver tissues by RT-PCR provided indirect evidence on the susceptibility of liver cells to SARS-CoV-2 infection [30]. This finding was supported by a recent report which showed the presence of SARS-CoV-2 viral particles in hepatocyte cytoplasm in two COVID-19 patients [20]. Indeed, Zhao et al recently found that SARS-CoV-2 infection impaired the barrier and bile acid transporting functions of cholangiocytes [23]. In our study, we further confirmed that human liver organoid derived intra-hepatic bile duct cells provided an excellent ex vivo model for studying SARS-CoV-2 infection. Cholangiocytes were found to co-express both ACE2 and TMPRSS2 and they were highly susceptible to coronavirus infection. One interesting finding was the identification of TMPRSS4 expression on these cells. In a recent study using an enteroid model, TMPRSS4 protein was shown to be another important serine protease which could work in synergy with TMPRSS2 to release SARS-CoV-2 into the host cell, after viral binding to ACE2 [31]. Thus, similar viral interaction and kinetics with intra-hepatic bile duct cells may be expected. Further experiments are underway to determine this.

The three highly pathogenic human coronaviruses include the severe acute respiratory syndrome coronavirus (SARS□CoV), the Middle East respiratory syndrome coronavirus (MERS□CoV) and the 2019 new coronavirus (SARS□CoV□2). These three viruses can cause diseases in various body systems. Similar to SARS-CoV-2, studies have shown that patients infected with SARS□CoV and MERS□CoV can also develop degrees of liver injury. As MERS-CoV uses dipeptidyl peptidase-4 (DPP4) as the spike protein receptor, as opposed to ACE2, we specifically studied and compared only SARS-CoV and SARS-CoV-2 here. Our data demonstrated that SARS-CoV-2 had much higher infectivity and replication rate than SARS-CoV on bile duct cells, which is similar to other studies on viral infectivity on respiratory and intestinal cells. These help explain why SARS-CoV-2 has caused such a huge worldwide pandemic over the past year. For the ability to infect the liver, previous histological studies showing lobular and portal activity in COVID-19 patients have not pinpointed at-risk cell populations, nor the underlying mechanisms in the course of liver injury [8,9]. Our finding of high susceptibility of intrahepatic cholangiocytes to SARS-CoV-2 infection suggests that biliary cholangiopathy cannot be overlooked both as a cause of liver injury and also as a potential source of viral shedding into the gastrointestinal tract. As COVID-19 pandemic continues to deteriorate, better understanding of SARS-CoV-2 infectivity on different cell populations is important in the development of comprehensive and targeted therapeutic and public health strategies.

## Data Availability

Data available upon request to corresponding authors

## Competing interests

All authors declare no support from any organization for the submitted work; no financial relationships with any organizations that might have an interest in the submitted work in the previous three years; no other relationships or activities that could appear to have influenced the submitted work.

## Author Contributions

Data contribution, Manuscript writing: V.L., K.H., H.Y., Data Analysis: K.H., R.B., M.C., Data contribution, Manuscript critique: P.C., P.T., Manuscript writing, Data coordination, Study Planning: V.L., M.C., K.W.

## Notes

### Competing Interest Statement

The authors have declared no competing interest.

### Funding Statement

There was no external funding for this study

### Author Declarations

Hong Kong West Cluster-Hong Kong University Cluster Research Ethics Committee/Institutional Review Board (UW 16-052)

